# Self-reported COVID-19 symptoms on Twitter: An analysis and a research resource

**DOI:** 10.1101/2020.04.16.20067421

**Authors:** Abeed Sarker, Sahithi Lakamana, Whitney Hogg-Bremer, Angel Xie, Mohammed Ali Al-Garadi, Yuan-Chi Yang

## Abstract

**Objective:** To mine Twitter to quantitatively analyze COVID-19 symptoms self-reported by users, compare symptom distributions against clinical studies, and create a symptom lexicon for the research community.

**Materials and methods:** We retrieved tweets using COVID-19-related keywords, and performed semi-automatic filtering to curate self-reports of positive-tested users. We extracted COVID-19-related symptoms mentioned by the users, mapped them to standard concept IDs (UMLS), and compared the distributions to those reported in early studies from clinical settings.

**Results:** We identified 203 positive-tested users who reported 1002 symptoms using 668 unique expressions. The most frequently-reported symptoms were fever/pyrexia (66.1%), cough (57.9%), body ache/pain (42.7%), fatigue (42.1%), headache (37.4%), and dyspnea (36.3%) amongst users who reported at least 1 symptom. Mild symptoms, such as anosmia (28.7%) and ageusia (28.1%) were frequently reported on Twitter, but not in clinical studies.

**Conclusion:** The spectrum of COVID-19 symptoms identified from Twitter may complement those identified in clinical settings.

## INTRODUCTION

The outbreak of the coronavirus disease 2019 (COVID-19) is one of the worst pandemics in the known World history.^1,2^ As of May 8, 2020, over 4 million confirmed positive cases have been reported globally, causing over 275,000 deaths.^3^ As the pandemic continues to ravage the world, numerous research studies are being conducted whose focuses range from trialing possible vaccines and predicting the trajectory of the outbreak to exploring the characteristics of the virus by studying those infected.

Early studies focusing on identifying the symptoms experienced by those infected by the virus mostly included patients who were hospitalized or received clinical care.^4–6^ Many infected people only experience mild symptoms or are asymptomatic and do not seek clinical care, although the specific portion of asymptomatic carriers is unknown.^7–9^ To better understand the full spectrum of symptoms experienced by infected people, there is a need to look beyond hospital- or clinic-focused studies. With this in mind, we explored the possibility of using social media, namely Twitter, to study symptoms self-reported by users who tested positive for COVID-19. Our primary goals were to (i) verify that users report their experiences with COVID-19—including their positive test results and symptoms experienced—on Twitter, and (ii) compare the distribution of self-reported symptoms with those reported in studies conducted in clinical settings. Our secondary objectives were to (i) create a COVID-19 symptom corpus that captures the multitude of ways in which users express symptoms so that natural language processing (NLP) systems may be developed for automated symptom detection, and (ii) collect a cohort of COVID-19-positive Twitter users whose longitudinal self-reported information may be studied in the future. To the best of our knowledge, this is the first study that focuses on extracting COVID-19 symptoms from public social media. We have made the symptom corpus public with this paper to assist the research community, and it will be part of a larger, maintained data resource—a social media COVID-19 Data Bundle (https://sarkerlab.org/covid_sm_data_bundle/).

## MATERIALS AND METHODS

### Data collection and user selection

We collected tweets, including texts and metadata, from Twitter via its public streaming application programming interface (API). First, we used a set of keywords/phrases related to the coronavirus to detect tweets through the API: *covid, covid19, covid-19, coronavirus*, and *corona AND virus*, including their hashtag equivalents *(e.g., #covid19)*. Due to the high global interest on this topic, these keywords retrieved very large numbers of tweets. Therefore, we applied a first level of filtering to only keep tweets that also mentioned at least one of the following terms: *positive, negative, test* and *tested*, along with at least one of the personal pronouns *I, my, us, we*, and *me*, and only these tweets were stored in our database. To discover users who self-reported positive COVID-19 tests, we applied another layer of filtering using regular expressions. We used the expressions ‘*i.*test[ed] positive*’, ‘*we.*test[ed’] positive*’, ‘*test.*came back positive*’, ‘ *my. *[covid\ coronavirus* | *covid19].*symptoms*’, and ‘*[covid\coronavirus\covid19].*[test\tested].*us’*. We also collected tweets from a publicly available Twitter dataset that contained IDs of over 100 million COVID-19-related tweets,^10^ and applied the same layers of filers. Three authors manually reviewed the tweets and profiles to identify true self-reports, while discarding the clear false positives *(e.g*., ‘*I dreamt that I tested positive for covid*… ’). We further removed users from our COVID-19-positive set if their self-reports were deemed to be fake or were duplicates of posts from other users, or if they stated that their tests had come back negative despite their initial beliefs about contracting the virus. These multiple layers of filtering gave us a manageable set of potential COVID-19-positive users (a few hundred) whose tweets we could analyze semi-automatically. The filtering decisions were made iteratively by collecting sample data for hours and days, and then updating the collection strategy based on analyses of the collected data.

### Symptom discovery from user posts

For all the COVID-19-positive users identified, we collected all their past posts dating back to February 1, 2020. We excluded non-English tweets and those posted earlier than the mentioned date. We assumed that symptoms posted prior to February 1 were unlikely to be related to COVID-19, particularly because our data collection started in late February and most of the positive test announcements we detected were from late March to early April. Since we were interested only in identifying patient-reported symptoms in this study, we attempted to shortlist tweets that were likely to mention symptoms. To perform this, we first created a meta-lexicon by combining MedDRA,^11^ Consumer Health Vocabulary (CHV),^12^ and SIDER.^13^ Lexicon-based approaches are known to have low recall particularly for social media data since social media expressions are often non-standard and contain misspellings.^14,15^ Therefore, instead of searching the tweets for exact expressions from the tweets, we performed inexact matching using a string similarity metric. Specifically, for every symptom in the lexicon, we searched windows of sequences of characters in each tweet that had similarity values above a specific threshold. We used the Levenshtein ratio as the similarity metric, computed as 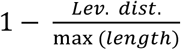, where *Lev. dist*. represents the Levenshtein distance between the two strings and *max(length)* represents the length of the longer string. Our intent was to attain high recall, so that we were unlikely to miss possible expressions of symptoms while also filtering out many tweets that were completely off topic. We set the threshold via trial and error over sample tweets, and because of the focus on high recall, this approach still retrieved many false positives (*e.g*., tweets mentioning body parts but not in the context of an illness or a symptom). After running this inexact matching approach on approximately 50 user profiles, we manually extracted the true positive expressions *(i.e*., those that expressed symptoms in the context of a COVID-19) and added them to the meta-lexicon.

Following these multiple filtering methods, we manually reviewed all the posts from all the users, identified each true symptom expressed, and removed the false positives. We semi-automatically mapped the expressions to standardized concept IDs in the Unified Medical Language System (UMLS) using the meta-lexicon we developed and the NCBO BioPortal.^16^ In the absence of exact matches, we searched the BioPortal to find the most appropriate mappings. Using Twitter’s web interface, we manually reviewed all the profiles, paying particularly close attention to those with less than five potential symptom-containing tweets, to identify possible false negatives left by the similarity based matching algorithms. All annotations and mappings were reviewed, and the reviewers’ questions were discussed at meetings. In general, we found that it was easy for annotators to detect expressions of symptoms, even when the expressions were non-standard *(‘pounding in my head* = Headache). Each detected symptom was reviewed by at least two authors, and the first author of the paper reviewed all the annotations.

Once the annotations were completed, we computed the frequencies of the patient-reported symptoms on Twitter and compared them with several other recent studies that used data from other sources. We also identified users who reported that they had tested positive and also specifically stated that they showed ‘no *symptoms’*. We excluded non-specific statements about symptoms, such as *‘feeling sick* and *‘signs of pneumonia*’. When computing the frequencies and percentages of symptoms, we used two models: (i) computing raw frequencies over all the detected users, and (ii) computing frequencies for only those users who reported at least 1 symptom or explicitly stated that they had no symptoms. We believe the frequency distribution for (ii) was more reliable as for users who reported no specific symptoms, we could not verify if they had actually experienced any symptoms and not reported them or just did not share symptoms over Twitter.

## RESULTS

Our initial keyword-based data collection and filtering from the different sources retrieved millions of tweets, excluding retweets. We found many duplicate tweets, which were mostly re-posts (not retweets) of tweets posted by celebrities. Removing duplicates left us with 305 users (499,601 tweets). 102 of them were labeled as *‘negatives*’—users who stated that their tests had come back negative, removed their original COVID-19-positive self-reports, or posted fake information about testing positive (*e.g*., we found some users claiming they tested positive as an April fool joke). This left us with 203 COVID-19-positive users with 68,318 tweets since February 1. The symptom detection approach reduced the number of unique tweets to review to 7,945.

The 203 users expressed 1002 total symptoms (mean: 4.94; median: 4) using 668 unique expressions, which we grouped into 46 categories, including a *‘No Symptoms’* category (Table 1). 171 users expressed at least 1 symptom or stated that they were asymptomatic (84.2%). 32 (15.8%) users did not mention any symptoms or only expressed generic symptoms, which we did not include in the counts (we provide these expressions in the lexicon accompanying this paper). 10 users explicitly mentioned that they experienced no symptoms. As Table 1 shows, *fever/pyrexia* was the most commonly reported symptom, followed by *cough, body ache & pain, headache, fatigue, dyspnea, chills, anosmia, ageusia, throat pain and chest pain*—each mentioned by over 20% of the users who reported at least one symptom. Figure 1 illustrates the first detected report of each symptom on a timeline, and Figure 2 shows the distribution of the number of symptoms reported by the cohort.

**Table 1.**
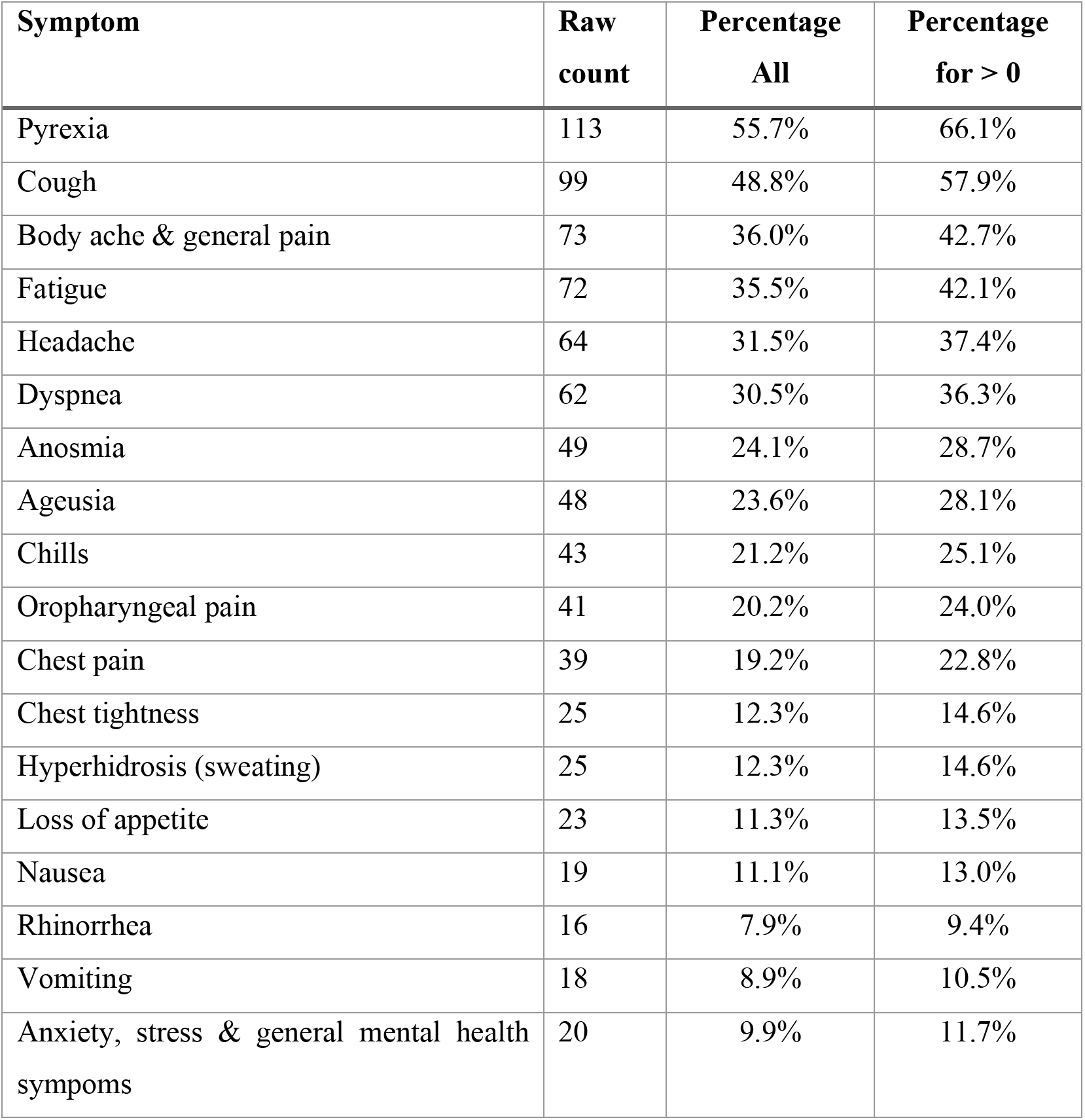
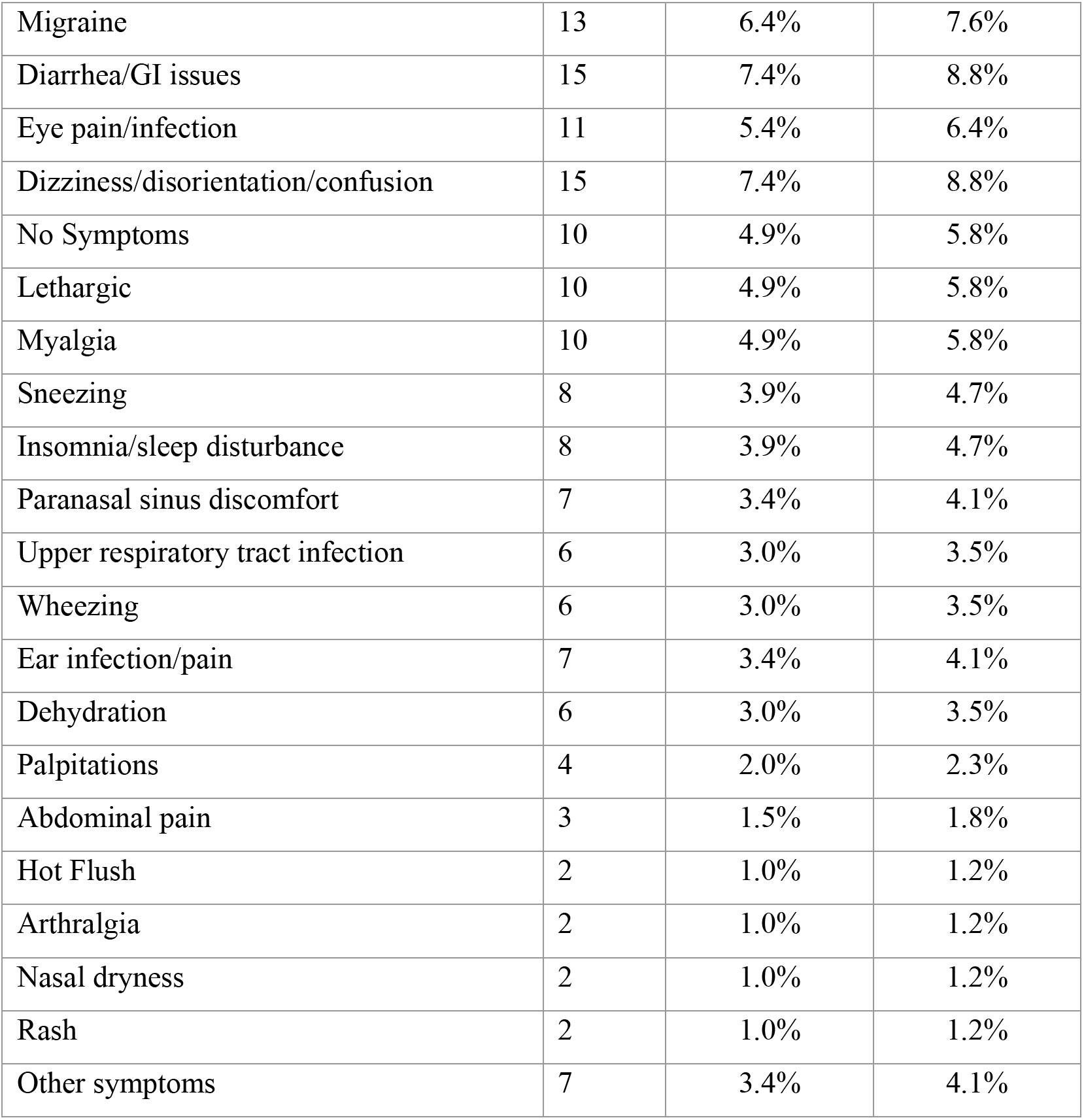
Distribution of symptoms reported by COVD-19 positive users on Twitter. Symptoms expressed once only are grouped under “Other symptoms”.

| Symptom | Raw count | Percentage All | Percentage for > 0 |
| --- | --- | --- | --- |
| Pyrexia | 113 | 55.7% | 66.1% |
| Cough | 99 | 48.8% | 57.9% |
| Body ache & general pain | 73 | 36.0% | 42.7% |
| Fatigue | 72 | 35.5% | 42.1% |
| Headache | 64 | 31.5% | 37.4% |
| Dyspnea | 62 | 30.5% | 36.3% |
| Anosmia | 49 | 24.1% | 28.7% |
| Ageusia | 48 | 23.6% | 28.1% |
| Chills | 43 | 21.2% | 25.1% |
| Oropharyngeal pain | 41 | 20.2% | 24.0% |
| Chest pain | 39 | 19.2% | 22.8% |
| Chest tightness | 25 | 12.3% | 14.6% |
| Hyperhidrosis (sweating) | 25 | 12.3% | 14.6% |
| Loss of appetite | 23 | 11.3% | 13.5% |
| Nausea | 19 | 11.1% | 13.0% |
| Rhinorrhea | 16 | 7.9% | 9.4% |
| Vomiting | 18 | 8.9% | 10.5% |
| Anxiety, stress & general mental health symptoms | 20 | 9.9% | 11.7% |
| Migraine | 13 | 6.4% | 7.6% |
| Diarrhea/GI issues | 15 | 7.4% | 8.8% |
| Eye pain/infection | 11 | 5.4% | 6.4% |
| Dizziness/disorientation/confusion | 15 | 7.4% | 8.8% |
| No Symptoms | 10 | 4.9% | 5.8% |
| Lethargic | 10 | 4.9% | 5.8% |
| Myalgia | 10 | 4.9% | 5.8% |
| Sneezing | 8 | 3.9% | 4.7% |
| Insomnia/sleep disturbance | 8 | 3.9% | 4.7% |
| Paranasal sinus discomfort | 7 | 3.4% | 4.1% |
| Upper respiratory tract infection | 6 | 3.0% | 3.5% |
| Wheezing | 6 | 3.0% | 3.5% |
| Ear infection/pain | 7 | 3.4% | 4.1% |
| Dehydration | 6 | 3.0% | 3.5% |
| Palpitations | 4 | 2.0% | 2.3% |
| Abdominal pain | 3 | 1.5% | 1.8% |
| Hot Flush | 2 | 1.0% | 1.2% |
| Arthralgia | 2 | 1.0% | 1.2% |
| Nasal dryness | 2 | 1.0% | 1.2% |
| Rash | 2 | 1.0% | 1.2% |
| Other symptoms | 7 | 3.4% | 4.1% |

**Figure 1.**
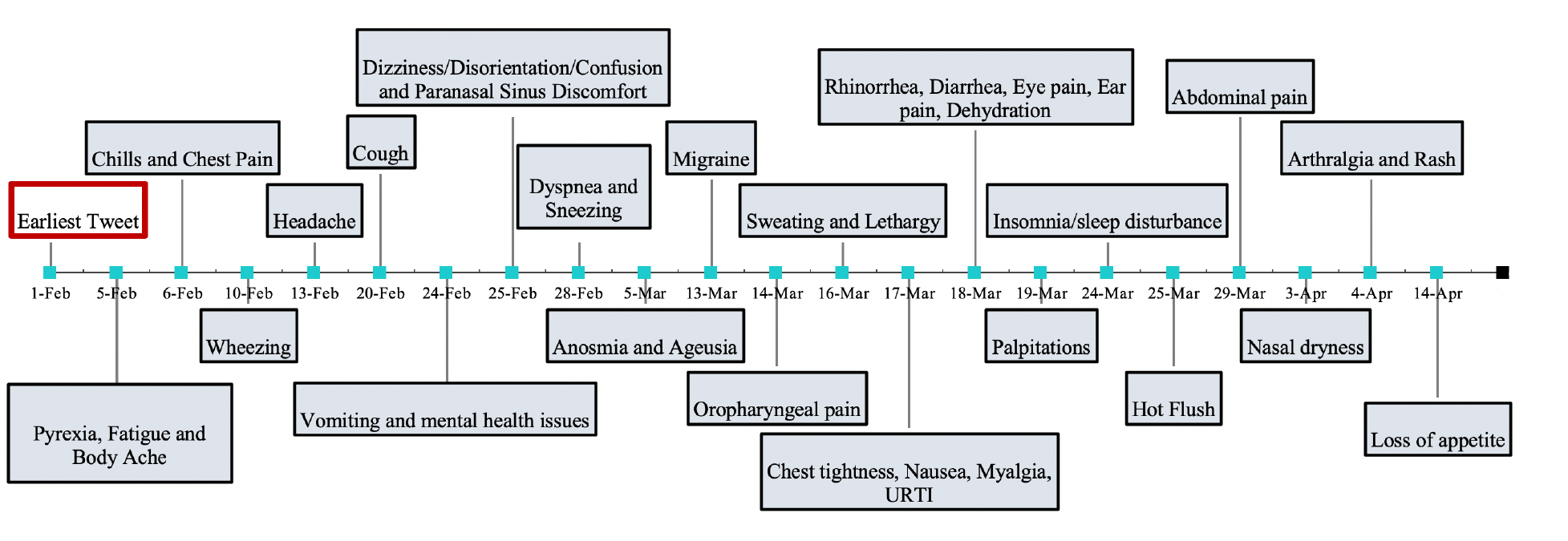
Timeline of first reports of each symptom by the Twitter cohort.

**Figure 2.**
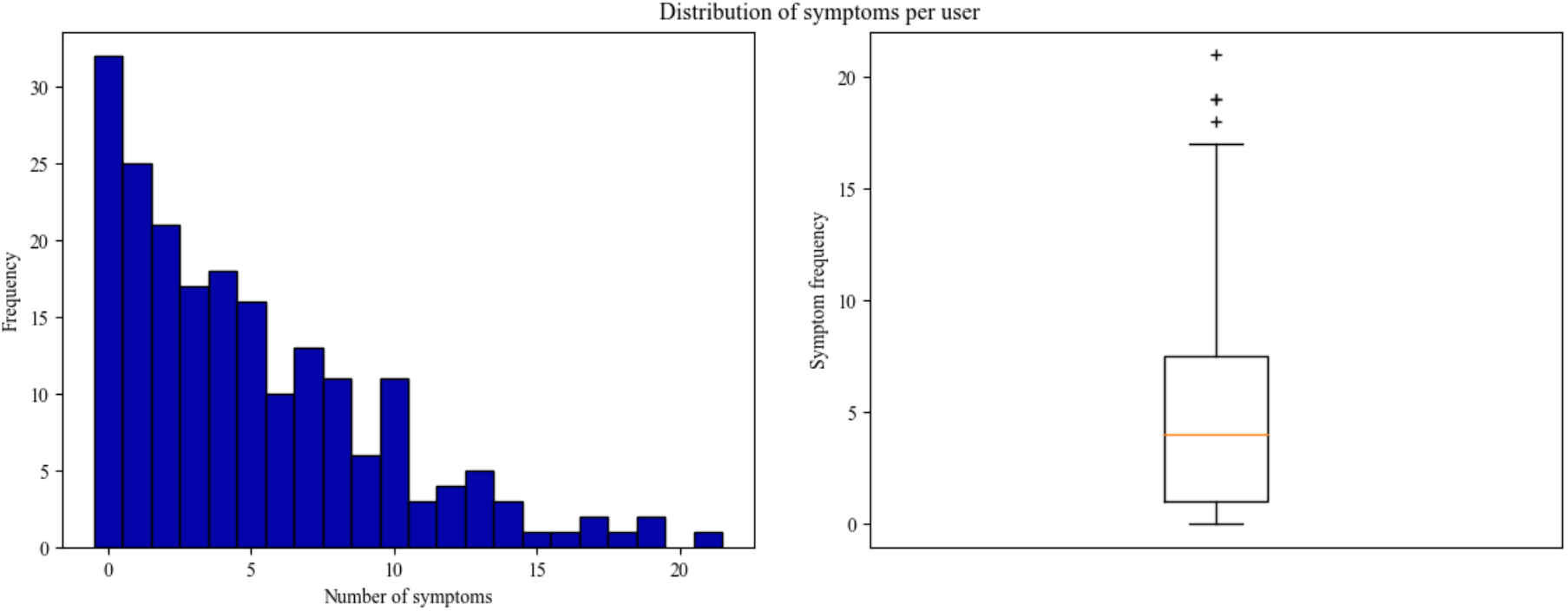
Distribution of the number of symptoms reported by the Twitter cohort.

**Table 2.** Comparison of common symptoms reported on Twitter by COVID-19-positive users with those reported in clinical settings.

| Symptom | Our study<sup>θ</sup><br>(n=169)<br>N (%) | Huang et al.<sup>6</sup> (n=41)<br>N (%) | Chen et al.<sup>5</sup><br>(n=249)<br>N (%) | Wang et al.<sup>17</sup><br>(n=138)<br>N (%) | Chen et al.<sup>18</sup> (n=99)<br>N (%) | Guan et al.<sup>4</sup><br>(n=1099)<br>N (%) | WHO Report<sup>19</sup><br>(n=55,924)<br>N (%) |
| --- | --- | --- | --- | --- | --- | --- | --- |
| Fever (Pyrexia) | 113 (66) | 40 (98) | 217 (87) | 136 (99) | 82 (83) | 975 (89) | 49157 (88) |
| Cough | 99 (58) | 31 (76) | 91 (37) | 82 (59) | 81 (82) | 745 (68) | 37861 (68)** |
| Dyspnea | 62 (36) | 22/40 (55) | 19 (8) | 43 (31) | 31 (31) | 205 (19) | 10402 (19) |
| Headache | 64 (37) | 3/38 (8) | 28 (11)<sup>∇</sup> | 9 (7) | 8 (8) | 150 (14) | 7606 (14) |
| Body ache & general pain | 73 (43) | - | - | 48 (35)<sup>ψ</sup> | 11 (11)<sup>ψ</sup> | 164 (15)<sup>ψ</sup> | 8277 (15)<sup>ψ</sup> |
| Fatigue | 72 (42) | 18 (44)* | 39 (16) | 96 (70) | - | 419 (38) | 21307 (38) |
| Chills | 43 (25) | - | - | - | - | 126 (12) | 6375 (11) |
| Anosmia | 49 (29) | - | - | - | - | - | - |
| Ageusia | 48 (28) | - | - | - | - | - | - |
| Chest pain | 39 (23) | - | - | - | 2 (2) |  | - |
| Oropharyngeal pain (sore throat) | 41 (24) | - | 16 (6) | 24 (17) | 5 (5) | 153 (14) | 7773 (14) |
| Diarrhea | 15 (9) | 1/38 (3) | 8 (3) | 14 (10) | 2 (2) | 42 (4) | 2069 (4) |
| Rhinorrhea | 16 (9) | - | 17 (7) | - | 4 (4) | 53 (5) | 2684 (5)<sup>ψψ</sup> |
| Anorexia | 23 (14) | - | 8 (3) | 55 (40) | - | - | - |
| Nausea | 19 (11) | - | - | 14 (10) | 1 (1)<sup>♣</sup> | 55 (5)<sup>♣</sup> | 2796 (5)<sup>♣</sup> |
| Asymptomatic | 10 (6) | - | 7 (3) | - | - | - | - |

<sup>θ</sup>For users who expressed at least 1 symptom or expressed that they did not have any symptoms.
\*The study provided a combined number for myalgia and fatigue.
<sup>∇</sup>Headache and dizziness was combined for this study.
<sup>ψ</sup>The reported number is for myalgia/muscle ache and/or arthralgia. In our study, we separated myalgia, arthralgia, body ache and pain.
<sup>♣</sup>Nausea and vomiting as a single category.
\*\*Reported as dry cough.
<sup>ψψ</sup>Reported as nasal congestion.

Table 2 compares the symptom percentages reported by our Twitter cohort with several early studies conducted in clinical settings *(i.e*., patients who were either hospitalized or visited hospitals/clinics for treatment). The top symptoms remained fairly consistent across the studies—*fever/pyrexia, cough, dyspnea, headache, body ache* and *fatigue*. The percentage of fever (66%), though the highest in our dataset, is lower than all the studies conducted in clinical settings. In our study, we distinguished, where possible, between *myalgia* and *arthralgia*, and combined *pain* (any pain other than those explicitly specified) and *body ache*. Combining all these into one category, as some studies had done, would result in a higher proportion. We found considerable numbers of reports of *anosmia* (29%) and *ageusia* (28%), with approximately one-fourth of our cohort reporting these symptoms. Reports of these symptoms, however, were missing from the referenced studies conducted in clinical settings.

## DISCUSSION AND CONCLUSIONS

Our study revealed that there were many self-reports of COVID-19 positive tests on Twitter, although such reports are buried in large amounts of noise. We observed a common trend among Twitter users of describing their day-to-day disease progression since the onset of symptoms. This trend perhaps became popular as celebrities started describing their symptoms on Twitter. We saw many reports from users who tested positive but initially showed no symptoms, and some who expressed anosmia and/or ageusia (first reported on March 5) as the only symptoms, which were undocumented in the comparison studies. There are some studies that suggest that anosmia and ageusia may be the only symptoms of COVID-19 among otherwise asymptomatic patients.^20–22^ The most likely explanation behind the differences between symptoms reported on Twitter and the clinical studies is that the former were reported mostly by users who had milder infections, while people who visited hospitals often went there to receive treatment for serious symptoms. Also, the median ages of the patients studied in clinical studies tended to be much higher than the median age of Twitter users (in the U.S., median Twitter user age is 40^23^). In contrast to the clinical studies, in our cohort, some users expressed mental health-related consequences (*e.g*., stress/anxiety) of testing positive. It was difficult in many cases to ascertain if the mental health issues were directly related to COVID-19 or whether the users had prior histories of such conditions.

To the best of our knowledge, this is the first study to have utilized Twitter to curate symptoms posted by COVID-19-positive users. In the interest of community-driven research, we have made the symptom lexicon available with this publication. The cohort of users detected over social media will enable us to conduct further studies in the future, enable us to study relatively unexplored topics such as the mental health impacts of the pandemic, and the long-term health of those infected by the virus.

## Data Availability

The symptom lexicon will be made available via the link provided.

https://sarkerlab.org/covid_sm_data_bundle/

## CONTRIBUTIONS

AS designed the study and data collection/filtering strategies. All authors contributed to the analyses, annotation process, and the writing of the manuscript.

## ACKNOWLEDGMENTS

TBA

